# Lexical and acoustic speech features relating to Alzheimer’s disease pathology

**DOI:** 10.1101/2021.09.27.21264148

**Authors:** Sunghye Cho, Katheryn A.Q. Cousins, Sanjana Shellikeri, Sharon Ash, David J. Irwin, Mark Liberman, Murray Grossman, Naomi Nevler

**Author notes:** Correspondence to:* Sunghye Cho, Linguistic Data Consortium, 3600 Market Street, Philadelphia, PA 19104.

## Abstract

**INTRODUCTION:** In this study, we compared digital speech features of AD and lvPPA patients in a biologically confirmed cohort and related them to specific neuropsychiatric test scores and CSF proteins.

**METHODS:** We extracted language variables with automated lexical and acoustic pipelines from oral picture descriptions of 44 AD and 21 lvPPA patients with autopsy or CSF confirmation of AD pathology. We correlated distinct speech features with MMSE and BNT test scores and CSF p-tau levels.

**RESULTS:** LvPPA patients produced fewer verbs, adjectives, and more fillers with lower lexical diversity and higher pause rate than AD. Both groups showed some shared language impairments compared with normal speakers.

**DISCUSSION:** Our speech measures captured differences in speech between the two phenotypes. Also, shared speech markers were linked to the common underlying pathology. This work demonstrates the potential of natural speech analysis in detecting underlying AD pathology.

## 1. Background

Speech production is a complex behavior, involving coordinated activation of multiple brain regions. Thus, examining speech production provides potential opportunities to identify neurodegenerative disease markers that are sensitive to specific phenotypes. Since Alzheimer’s disease (AD) accounts up to 80% of patients with dementia,^1^ much attention has been paid to cognitive and linguistic profiling of AD. Language produced by amnestic AD patients was found to be “empty” with abundance of nonspecific words, circumlocutions, and sparse content.^2^

Logopenic variant primary progressive aphasia (lvPPA) is a recently identified PPA variant,^3,4^ which is an atypical, non-amnestic manifestation of AD,^3,5–9^ with over 50% of autopsied cases associated with underlying AD pathology.^9,10^ Since the identification of this PPA variant, many studies have been dedicated to characterizing its linguistic features. Previous studies have shown that lvPPA patients speak slowly with impaired lexical access,^11^ and have poor phonemic discrimination^12^ with limited auditory-verbal short-term memory, naming impairment,^4,5^ and dysfluencies.^13^

Previous studies of neurodegenerative patients suggest that language features are useful as a screening tool^14–18^, because speech is easy to collect non-invasively, and it is sensitive to cognitive impairments. Despite the shared pathology of AD and lvPPA, previous studies have focused on the linguistic profiling of these two syndromes separately. With few comparative studies, this leaves an important gap in the literature. Most previous quantitative work has focused on measures such as the Boston Naming Test (BNT) that assess lexical retrieval during confrontation naming of an object. However, descriptions of natural, connected speech in AD and lvPPA are frequently informal. In this study, we identified similarities and differences between AD and lvPPA patients with biological confirmation of underlying AD pathology, by analyzing digitized, natural speech samples with reliable and reproducible automated methods. Based on previous studies, we hypothesized that lvPPA patients would produce more dysfluent speech with more limited lexical content than AD patients. We also hypothesized that AD and lvPPA patients would share linguistic similarities, including decreased speech production. We associated language variables with clinical test scores and CSF analytes for additional validation and specific mechanistic clarification.

## 2. Methods

### 2.1 Participants

We examined oral picture descriptions that were collected from 93 participants in the Department of Neurology at the Hospital of the University of Pennsylvania. Forty-four participants had amnestic AD, and 21 were lvPPA patients with confirmed underlying AD pathology, based on autopsy (n=15; 4 lvPPA and 11 AD patients) or on cerebrospinal fluid (CSF) analytes levels (n=50; 17 lvPPA and 33 AD patients; phosphorylated Tau (p-Tau)/beta-amyloid 42 (Aβ) ≥ 0.09^19^ *and* total Tau/Aβ ≥ 0.34^20^). Twenty-eight matched elderly healthy controls (HC) were included as a control group. Participants with other neurological, psychiatric, or medical conditions that could affect cognition were excluded.

Table 1 shows the demographic and clinical characteristics of the participants. The groups did not differ in age, sex, or education level. The patient groups did not differ from each other in disease duration (*p*=0.45), the Mini-Mental State Exam (MMSE; *p*=0.12), BNT (*p*=0.32), CSF p-Tau (*p*=0.81), Aβ (*p*=0.36), or p-Tau/Aβ ratio levels (*p*=0.66).

**Table 1:** Demographic and clinical characteristics of participants.

|  | AD (N=44) | lvPPA (N=21) | HC (N=28) | p value |
| --- | --- | --- | --- | --- |
| **Age (years)** | 62.0 (8.0) | 64.1 (8.2) | 65.9 (5.9) | 0.098 |
| **Education (years)** | 16.0 (2.6) | 16.1 (3.2) | 15.9 (2.6) | 0.965 |
| **Sex** |  |  |  | 0.986 |
| Female | 24 (54.5%) | 11 (52.4%) | 15 (53.6%) |  |
| Male | 20 (45.5%) | 10 (47.6%) | 13 (46.4%) |  |
| **Disease Duration (years)** | 3.7 (2.6) | 3.4 (1.5) | NA |  |
| **MMSE\*** | 21.1 (4.8) | 23.0 (4.2) | 29.2 (1.0) | < 0.001 |
| **BNT\*** | 20.1 (8.4) | 18.6 (8.6) | 27.9 (2.7) | < 0.001 |
| **CSF p-Tau** | 38.5 (20.1) | 38.4 (16.0) | NA |  |
| **CSF Aβ** | 139.9 (36.2) | 135.0 (26.2) | 415.0 (NA) | < 0.001 |
| **CSF p-Tau/Aβ** | 0.3 (0.2) | 0.3 (0.2) | NA |  |

NOTE: \*AD and lvPPA patients did not differ in MMSE or BNT.
Abbreviations: AD = Alzheimer’s disease, lvPPA = logopenic variant primary progressive aphasia, HC = healthy controls; MMSE = Mini-mental State Exam; BNT = Boston Naming Test; CSF = Cerebrospinal fluid; p-Tau = phosphorylated Tau; Aβ = beta-amyloid 42.

### 2.2 Data collection

To optimize reliability and reproducibility, we digitally recorded the participants’ descriptions of the Cookie Theft picture from the Boston Diagnostic Aphasia Examination.^21^ Descriptions were about 1 minute long. Recordings were orthographically transcribed. The earliest recording of each participant was analyzed with our lexical and acoustic pipelines, as described below.

### 2.3 Lexical pipeline

We automatically tagged the part-of-speech (POS) category of all tokens, using spaCy^22^ with its large language model (‘en_core_web_lg’) for English. The number of tense-inflected verbs was calculated by summing the number of modal auxiliaries, past-tense, and present-tense verbs. Dysfluency markers, including fillers, repetitions, and partial words, were counted separately. The count of each POS category, tense-inflected verbs, and dysfluency markers were converted to counts per 100 words, controlling for the total number of words per participant.

We rated each word for concreteness,^23^ semantic ambiguity,^24^ frequency,^25^ age of acquisition (AoA),^26^ familiarity,^26^ and word length by the number of phonemes,^27^ based on published norms. We calculated the mean scores of these measures for content words (nouns, verbs, adjectives, and adverbs) per participant.

Lastly, we measured lexical diversity, using the moving-average type-token ratio (MATTR),^28^ which has been described as one of the most reliable measures for calculating lexical diversity.^29^ The window length was set at 15 words. Detailed description of the lexical pipeline and validation of the POS tagging accuracy has been previously published.^30^

### 2.4 Acoustic pipeline

We employed an in-house speech activity detector (SAD) to segment audio recordings into speech segments and silent pauses. We visually reviewed the segments, and non-speech segments at the beginning and end of each recording and interviewer’s prompts were excluded from the analysis.

Using the SAD output, we calculated five duration-related measurements, including mean speech segment duration, mean pause segment duration, total speech time, percent of speech, and pause rate per minute. We summed the number of syllables of all words from the published norm^27^ and computed articulation rate as the number of syllables per second.

Additionally, we pitch-tracked all speech segments with Praat.^31^ To normalize physiological differences in pitch (f0), we converted the pitch values from Hz to semitones (st) using the 10th pitch percentile of each participant as a baseline: st=12*log_2_(f0/baseline f0). We used the converted 90th percentile as a measure of the pitch range of each speaker. Detailed description of the acoustic pipeline has been published previously,^32^ and the list of all analyzed features is included in the Appendix.

### 2.5 CSF analysis

Forty-two (30 AD and 12 lvPPA) patients had CSF biomarkers collected within one year of the Cookie Theft recording (mean interval = 4.7 months ± 3.5), including A*β* and p-Tau. This subset did not differ demographically or clinically from the larger group of patients. CSF was analyzed with two platforms, Luminex xMAP or Innotest ELISA, which was then transformed to the Luminex scale.^33^ We previously related CSF p-Tau levels directly to cerebral tau burden in our autopsy cohort.^33^ The AD and lvPPA subset groups did not differ in age (*p*=0.64), sex (*p*=0.99), education (*p*=0.82), disease duration (*p*=0.75), or the time difference between CSF sample collection and the Cookie Theft recording (*p*=0.43). To determine the association of language features with in vivo measures of pathology, we examined the relationship between our language variables and the two CSF biomarkers of Aβ and p-Tau.

### 2.6 Statistical considerations

To compare the groups, we tested if requirements for parametric tests were met with a Levene’s test. If the data met the requirements for parametric tests, we performed an Analysis of Variance. If not, we performed a Kruskal-Wallis test. We visually assessed residuals of the models to make sure the data was suitable for linear modelling. When a group difference was significant, we additionally performed a posthoc pairwise t-test or pairwise Wilcoxon rank sum test for pairwise group comparisons, adjusting *p*-values for multiple group comparisons (n=3) with the false discovery rate (fdr). We reported the effect size of each group comparison using Cohen’s *d*.

Patients’ language variables were z-scored using HC’s mean and standard deviation. These z-scores were used for visualization and linear regressions to estimate relations of our language variables and clinical ratings. We did not use z-scores to determine significant group differences with Z-tests, since the test statistic in some variables did not follow a normal distribution.

The z-scored language variables that showed significant group differences were associated with patients’ MMSE and BNT scores to investigate the relations of our language features to clinical ratings of cognitive and language impairment. To examine potential interactions, we included phenotype as an interaction term (MMSE/BNT ∼ language variable*phenotype). *P*-values were adjusted with fdr.

To validate our findings with levels of specific CSF biomarkers, we correlated patients’ CSF analyte levels to the language variables using Pearson correlation tests. CSF p-Tau levels were log-transformed to normalize the data. We also checked if patients’ clinical phenotype and the time difference between Cookie Theft recording and CSF sample collection were significant factors with linear regression models. Since the two factors were not significant, we only reported the results of simple correlations to simplify the models. All statistical analyses were performed with R version 4.1.0^34^ and RStudio version 1.4.1717.^35^

### 2.7 Ethics

The Institutional Review Board of the Hospital of the University of Pennsylvania approved the study of human subjects, and all participants agreed to participate in the study by written consent. All digital data was stored in secured HIPAA-compliant servers and handled by personnel trained in PPI protection.

## 3. Results

### 3.1 Differences between amnestic AD and lvPPA patients

LvPPA patients produced fewer tense-inflected verbs compared to AD (*p*=0.001, |*d*|=0.94) and HC (*p*=0.048, |*d*|=0.61; Fig.1A). AD patients’ tense-inflected verb counts did not significantly differ from HC (*p*=0.124, |*d*|=0.4). Patients with lvPPA showed lower lexical diversity than AD (*p*=0.05, |*d*|=0.52) and HC (*p*=0.005, |*d*|=1.02), yet AD patients did not differ from HC (*p*=0.149, |*d*|=0.39; Fig.1B). LvPPA patients produced fewer adjectives than AD patients (*p*=0.019, |*d*|=0.66) and HC (*p*<0.001, |*d*|=1.72); AD patients also produced fewer adjectives than HC (*p*=0.003, |*d*|=0.75; Fig.1C). Thus, both patient groups were impaired in their adjective production, but lvPPA was more severely impaired compared with AD.

**Figure 1:**
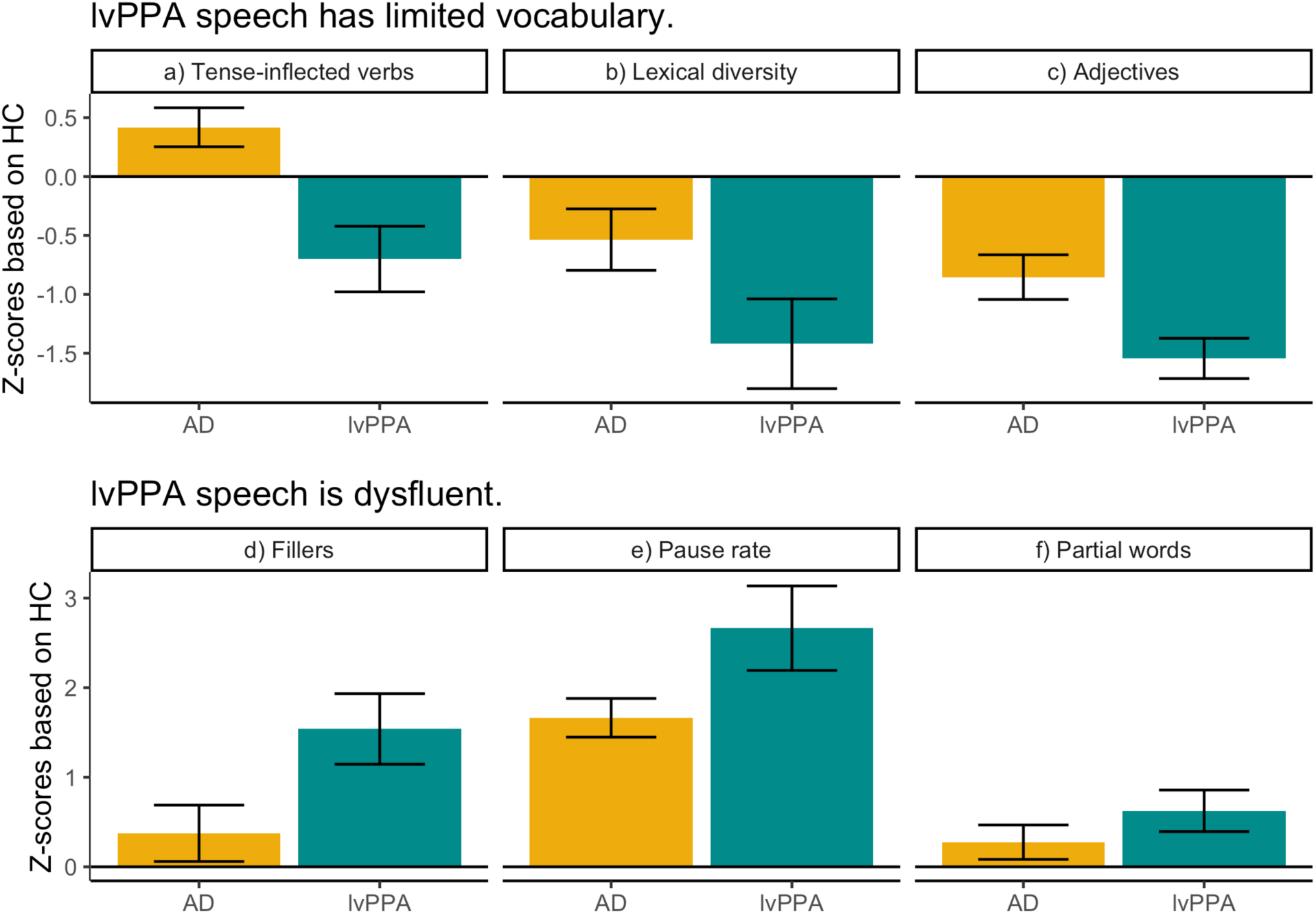
Speech differences between lvPPA and amnestic AD. For ease of visualization, z-scored values compared to HC mean and standard deviation are plotted. Horizontal black line indicates HC’s mean. Abbreviations: AD = Alzheimer’s disease, lvPPA = logopenic variant primary progressive aphasia, HC = healthy controls

LvPPA patients also produced more fillers than AD (*p*=0.022, |*d*|=0.6) and HC (*p*=0.01, |*d*|=1.06; Fig.1D), while AD did not significantly differ from HC (*p*=0.383, |*d*|=0.23). Patients with lvPPA showed a higher pause rate than AD (*p*=0.015, |*d*|=0.55) and HC (*p*<0.001, |*d*|=1.58; Fig.1E); AD patients’ pause rate was also higher than HC (*p*<0.001, |*d*|=1.34). Lastly, lvPPA patients produced more partial words than HC (*p*=0.042, |*d*|=0.61), but lvPPA patients did not significantly differ from AD speakers(*p*=0.147, |*d*|=0.3); AD patients did not differ from HC (*p*=0.313, |*d*|=0.24). Thus, lvPPA speakers produced an abnormal number of partial words, while AD speakers did not.

### 3.2 Impaired speech features in both AD and lvPPA

**Figure 2:**
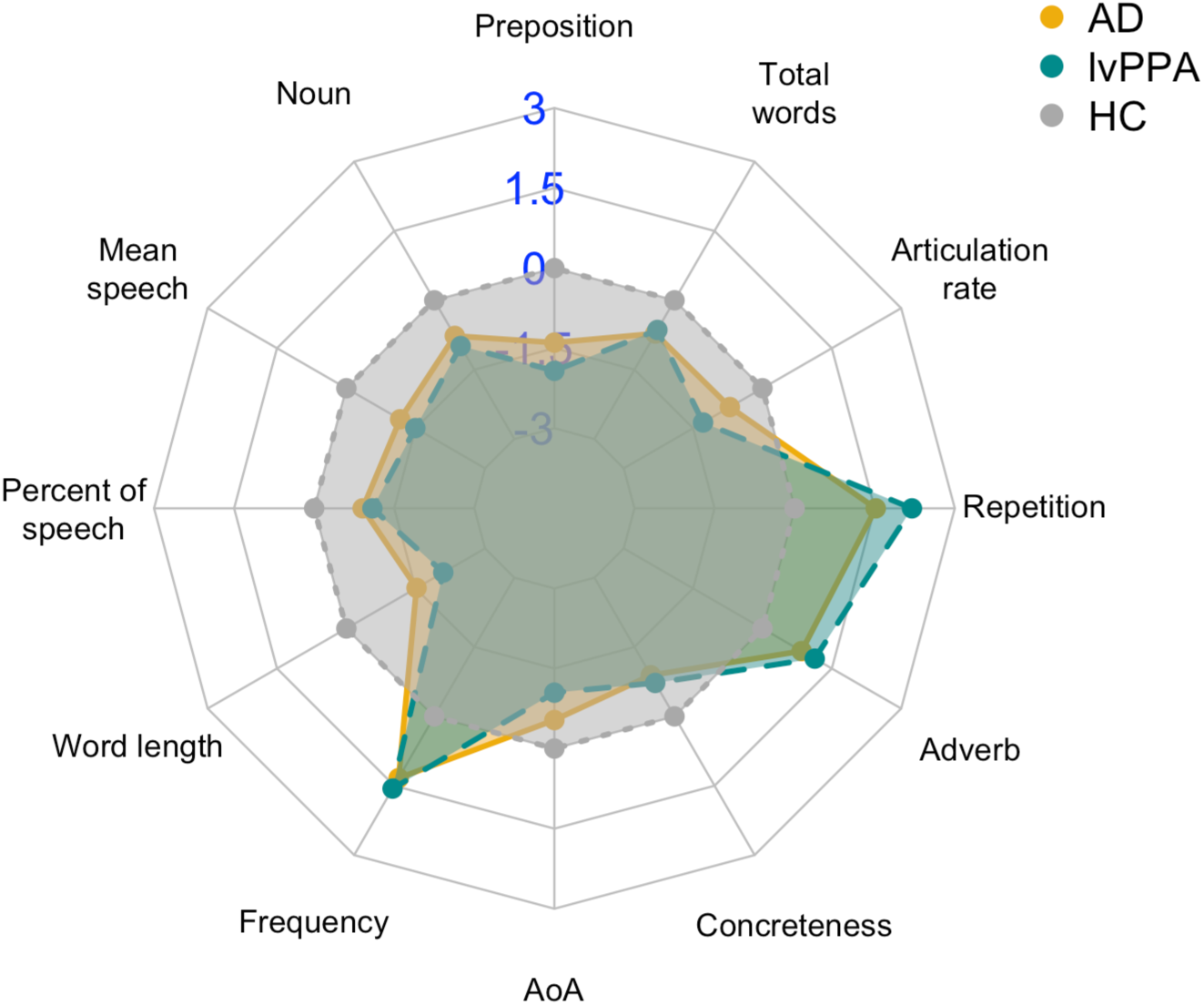
Impaired speech measures in both lvPPA (green) and AD (yellow). Only speech measures that were significantly different from HC (gray) are plotted, and measures were standardized based on the HC mean and standard deviation. The numbers in blue indicate z-scored values based on HC. Abbreviations: AD = Alzheimer’s disease, lvPPA = logopenic variant primary progressive aphasia, HC = healthy controls

AD and lvPPA patients produced fewer prepositions and nouns than HC; patients produced shorter speech segments than HC, and their percent of speech time out of the total time was also lower than HC (*p*<0.001, |*d*|>0.8 for all comparisons). Both groups’ content words were shorter, more frequent (length & frequency: *p*<0.001, |*d*|>0.8), earlier-acquired (lvPPA: *p*<0.001, |*d*|=0.95; AD: *p*<0.001, |*d*|=0.56), and more concrete (lvPPA: *p*=0.028, |*d*|=0.7, AD: *p*=0.002, |*d*|=0.86) than HC’s. Both patient groups produced more adverbs (lvPPA: *p*=0.029, |*d*|=0.84; AD: *p*=0.029, |*d*|=0.73) and repetitions (lvPPA: *p*<0.001, |*d*|=1.39; AD: *p*<0.001, |*d*|=0.79) than HC. Patients also spoke slower (lvPPA: *p*<0.001, |*d*|=1.01; AD: *p*<0.001, |*d*|=0.57), and they produced fewer words in total than HC (lvPPA: *p*=0.032, |*d*|=0.7; AD: *p*=0.007, |*d*|=0.73).

### 3.3 Relationships to clinical measures

**Figure 3:**
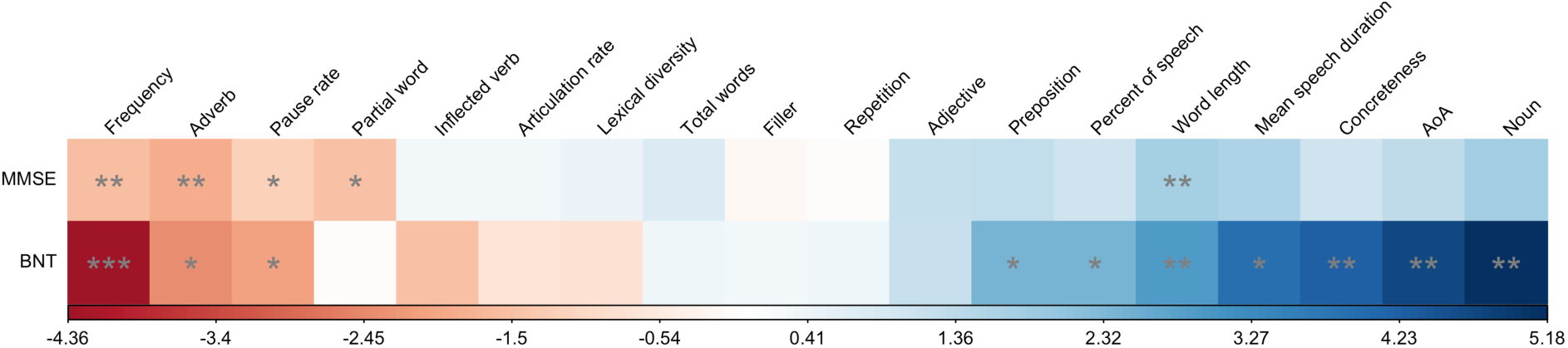
Results of linear regression models of speech masures in patients with MMSE and BNT. Standardized estimated coefficients (x-axis) are plotted with colors: positive coefficients are in blue and negative ones in red. P-values are adjusted with fdr. *: *p*<0.05, **: *p*<0.01, ***: *p*<0.001. Abbreviations: AD = Alzheimer’s disease, lvPPA = logopenic variant primary progressive aphasia, HC = healthy controls, MMSE = Mini-mental State Exam; BNT = Boston Naming Test

MMSE was significantly related to five language variables. Patients with lower MMSE produced more frequent (*β*=-1.6, *p*=0.009) and shorter content words (*β*=1.75, *p*=0.001), paused more frequently (*β*=-1.21, *p*=0.041), and produced more adverbs (*β*=-1.9, *p*=0.003) and partial words (*β*=-1.52, *p*=0.021).

BNT was significantly associated with ten variables. Patients with lower BNT produced frequent words (*β*=-4.36, *p*<0.001), many adverbs (*β*=-2.41, *p*=0.041), and had a high pause rate (*β*=-2.09, *p*=0.041). Patients with lower BNT also produced fewer prepositions (*β*=2.42, *p*=0.048) and nouns (*β*=5.18, *p*=0.003). Patients with lower BNT showed a low percent of speech produced during the picture description (*β*=2.4, *p*=0.045), and they produced shorter (*β*=2.94, *p*=0.003), less concrete (*β*=4.24, *p*=0.003), and earlier-acquired (*β*=4.69, *p*=0.003)content words, with shorter speech segments (*β*=3.89, *p*=0.041).

Only one variable showed a significant interaction with phenotype. AD patients with lower MMSE scores produced more adverbs (*β*=-1.9, *p*=0.003), yet lvPPA patients with lower MMSE scores produced fewer adverbs (*β*=2.67, *p*=0.015).

### 3.4 CSF results

**Figure 4:**
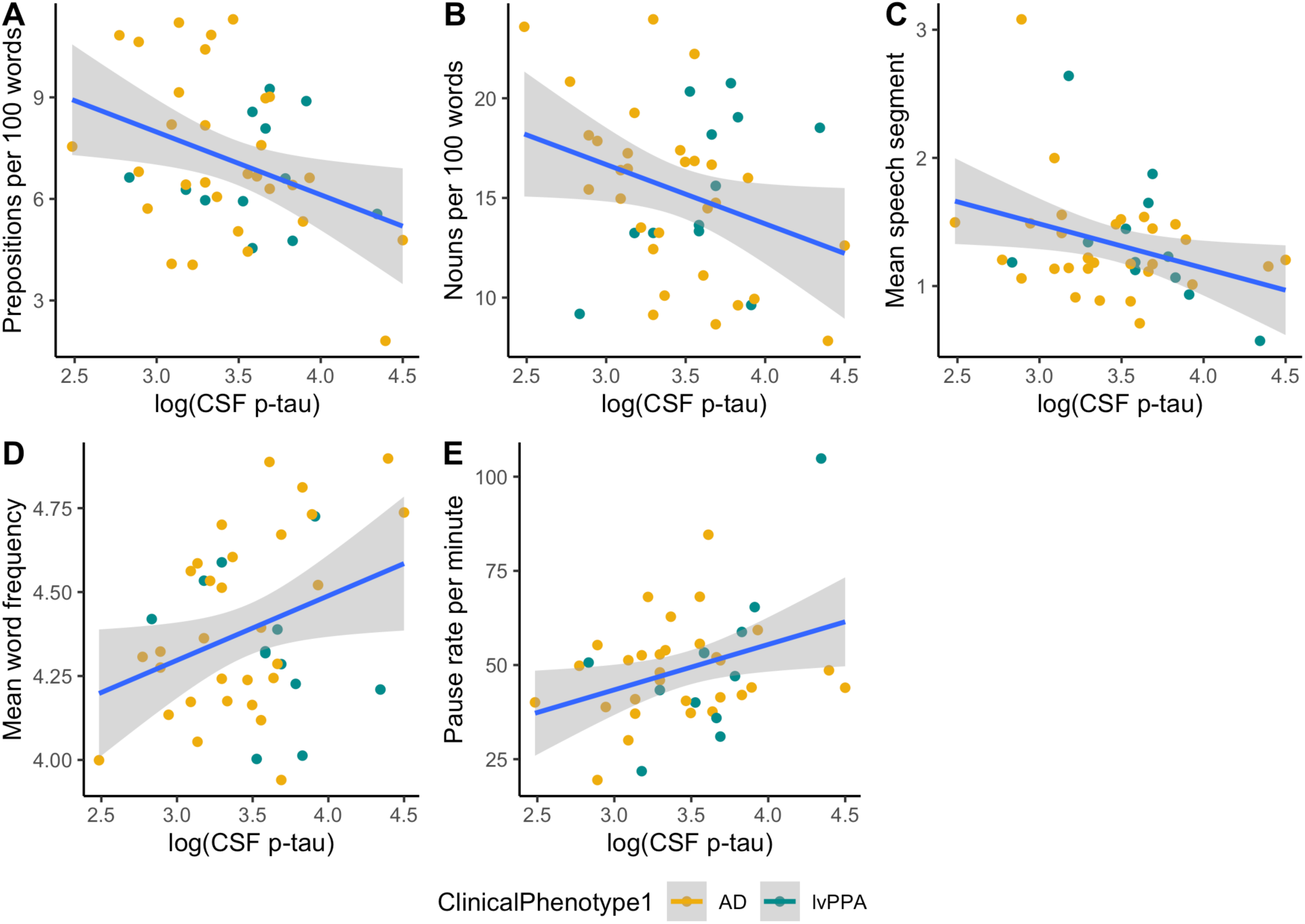
Significant correlations of the speech variables and CSF p-Tau levels. Abbreviations: AD = Alzheimer’s disease, lvPPA = logopenic variant primary progressive aphasia, HC = healthy controls, CSF = Cerebrospinal fluid, p-Tau = phosphorylated Tau, Aβ = beta-amyloid 42 Patients’ CSF p-Tau level correlated with lower preposition counts (*r*=-0.36, *p*=0.019), lower noun counts (*r*=-0.31, *p*=0.047), and shorter mean speech segment durations (*r*=-0.33, *p*=0.032; Fig.4A-C). Also, patients’ p-Tau levels were inversely correlated with a higher pause rate (*r*=0.34, *p*=0.026) and higher word frequency (*r*=0.33, *p*=0.036; Fig.4D-E). The other measures (adverbs, partial words, tense-inflected verbs, articulation rate, lexical diversity, total words, fillers, repetitions, adjectives, percent of speech, word length, concreteness, AoA) were not related to patients’ p-Tau levels. Aβ alone did not significantly correlate with any of the language measures.

## 4. Discussion

LvPPA is most frequently associated with underlying AD pathology, but direct comparison of lvPPA with amnestic AD patients has been reported rarely. The current study focuses on characterizing the language similarities and differences between lvPPA and AD in a biologically confirmed cohort. To optimize reliability and reproducibility, we used fully automated lexical and acoustic analyses to characterize linguistic markers of AD pathology. We expected that lvPPA patients with non-amnestic AD would produce more dysfluent speech with limited vocabulary than amnestic AD patients because of lvPPA’s phenotypic characteristics. Results confirmed that lvPPA patients produced fewer adjectives and tense-inflected verbs with lower lexical diversity than amnestic AD patients and HC. lvPPA patients also paused more frequently and produced more fillers and partial words than HC and/or amnestic AD patients. However, we also found that both patient groups shared impairments in some speech features relative to HC. For example, both patient groups produced more adverbs, including words like “*there*” and “*here*,” but fewer prepositions and nouns than HC. Also, patients’ content words were earlier-acquired, shorter, more frequent, and less concrete than those of HC. Patients produced more repetitions and fewer total words with a slower articulation rate and a shorter mean speech duration than HC. Some of these language variables were significantly related to clinical test scores and p-Tau levels in CSF. We discuss these findings below.

The patient groups significantly differed on six language measures: pause rate, partial words, fillers, adjectives, tense-inflected verbs, and lexical diversity. Fillers, adjectives, tense-inflected verbs, and lexical diversity were significantly more impaired in lvPPA than AD patients, emphasizing the deficits in lexical retrieval and poor fluency in these patients. However, these linguistic features distinguishing between lvPPA and amnestic AD were not related to patients’ MMSE or BNT. It is important to note that the mechanism thought to subserve retrieval of a single word in response to a stimulus picture appears to differ from lexical retrieval during natural connected speech.^36^ It is thus critical to monitor these speech features since they are not easily explained by more general and commonly used measures such BNT and MMSE. To our knowledge, the result that tense-inflected verb production differed between lvPPA and AD has not been previously reported. This finding seems to suggest that lvPPA patients produced fewer complete sentences, assuming that there was one tense-inflected verb per inflection phrase.^37^ This would be consistent with the observation that lvPPA patients’ limited auditory-verbal short-term memory results in briefer sentences.^3,4^ Frequent fillers in lvPPA were in line with previous observations, and consistent with their limited lexical retrieval.^11,38,39^ Lower adjective counts in amnestic AD patients compared to HC have been previously reported,^40^ yet we further showed that the adjective production was more impaired in non-amnestic lvPPA than amnestic AD. Lexical diversity has been frequently examined in the AD literature,^14,15,41,42^ where previous studies have found that AD patients’ lexical diversity was lower than that of HC. We showed that lexical diversity was even lower in lvPPA than in AD. The fact that lvPPA and AD patients significantly differed on these measures suggests that our language variables may capture subtle but unique phenotypic differences between lvPPA and AD. Additionally, traditional clinical ratings such as the MMSE and BNT are relatively insensitive to these linguistic features. Also, none of the six variables, except pause rate, correlated with CSF p-tau, and regression analyses failed to find an interaction of p-Tau level with phenotype in the assessment of pause rate. This suggests that these speech markers are related more narrowly to the phenotype and not necessarily to the underlying AD pathology burden. Further studies, including the anatomic distribution of pathology in an autopsy cohort with quantitative measures of pathological burden, may help shed light on this issue.

Some speech measures were related to MMSE and BNT. Pause rate showed more impairment in lvPPA than AD. This might indicate word-finding difficulty in lvPPA which could provoke frequent pausing to recall an appropriate word from their lexicon. It could also be that lvPPA patients spoke slowly – patients’ articulation rate was lower than that of HC – due to their difficulty in retrieving words to generate utterances. Pause rate was significantly related to both MMSE, an indicator of general cognitive impairment, and BNT, a measure of confrontation naming. Elevated pause rate, therefore, may reflect in part both patients’ word-finding difficulties and general cognitive impairments. Partial word count, which was impaired only in lvPPA, only correlated with MMSE, suggesting that it reflected in part lvPPA patients’ disease severity and general cognitive impairments, but not impaired object naming.

Word-finding difficulty in AD and lvPPA has been previously noted,^11,43–48^ where studies have shown that patients had impairments in auditory-verbal short-term memory and could not recall the phonological form of a word. However, comparative studies have not been reported to examine if both amnestic AD and non-amnestic lvPPA patients would show word-finding difficulty to a similar degree during natural speech. In our study, both patient groups produced content words that were more abstract, earlier-acquired, more frequent and shorter than those of HC, suggesting that they had difficulties in retrieving the full spectrum of lexical items needed to describe the picture. Word frequency and length were significantly related to both MMSE and BNT, which suggests that these lexical measures reflect in part patients’ disease severity and difficulties in lexical retrieval during confrontation naming. On the other hand, concreteness and AoA were significantly associated only with BNT, indicating that these may be more sensitive to word-finding difficulty in patients. AD and lvPPA patients did not significantly differ in these measures, confirming that some degree of word-finding difficulty is present in both amnestic and non-amnestic AD.

Adverb counts were greater in patients compared to HC. This may be related to patients frequently using “pro-adverbs,” including “*here*” and “*there*,” which replaced locational prepositional phrases. Patients typically produced utterances like “Mom is standing *here*,” for example, when HC produced “Mom is standing *in front of the sink*.” Elevated adverb counts were associated with low BNT scores in AD, suggesting that greater adverb use reflected AD patients’ difficulties in naming specific locations during natural speech. Patients’ difficulty in producing locational phrases were also partly reflected in the decreased preposition counts compared to HC, which was also related to BNT scores. Additional work is needed to determine if decrease preposition counts and increase adverb use are related to temporal propositions and other features in connected speech.

Some of our language variables correlated with CSF p-Tau levels, but not with A*β*. This finding is in line with previous findings that patients’ cognitive impairment is generally not related to A*β* levels but to accumulation of p-Tau.^49–51^ Language production is one of the most essential daily functions of humans, which needs to be taken into consideration in AD clinical trials and may serve in monitoring response to treatment. Since our automated procedures for collecting speech features is highly reliable and reproducible, investigations of speech variables as secondary outcome measures should be considered in disease-modifying trials targeting tau.

The strengths of our study include that we directly compared speech features in AD and lvPPA patients with biological evidence of underlying AD pathology. We examined natural connected speech quantitatively using automated analyses of digitized speech samples. We inspected language differences and similarities in these groups and showed that our language variables could capture subtle linguistic differences between the two phenotypes, and traditional cognitive measures such as MMSE and BNT appear to be insensitive to some of the features distinguishing between amnestic AD and lvPPA. These methods may be useful in screening for underlying AD pathology and monitoring disease progression and response to therapeutic interventions, because collecting one-minute speech samples is easy, highly reproducible, and less costly and can be collected remotely compared with other biomarkers. Future studies, testing the value of these speech features in longitudinal datasets and developing machine learning classifiers for patient stratification of AD pathology would be valuable. Shortcomings include the assessment of relatively small samples, the difficulty obtaining train-test generalizability data in rare lvPPA and early-onset AD cases such as these, the use of a single stimulus picture to elicit the speech sample, and the absence of high resolution MRI data to assess the anatomic associates of these linguistic features.

## 5. Conclusion

We implemented automated methods in analyzing acoustic and lexical characteristics of amnestic AD and lvPPA patients’ natural speech.We found speech markers that differed between the groups. These features seemed to relate to the clinical phenotype and less so to the underlying pathology. We also found speech markers that were shared between these two AD phenotypes and linked to the common underlying pathology. This work demonstrates the potential of natural speech to reflect underlying AD pathology while distinguishing between specific phenotypes with the same pathology. Considering the cost effectiveness and reliability of speech data, such markers could contribute to screening in AD clinical trials and longitudinal monitoring in a more precise and inclusive way.

## Data Availability

Data will be available upon request from appropriate research groups.

## Acknowledgements

The authors thank the patients and caregivers who participated in this study. Funding: This study was funded by grants from the National Institute of Health (AG066597, AG054519, NS109260, P30 AG072979), Alzheimer’s Association (AACSF-18-567131, AARF-D-619473, AARF-D-619473-RAPID), Department of Defense (PR192041).

## Appendix

**Table A.**
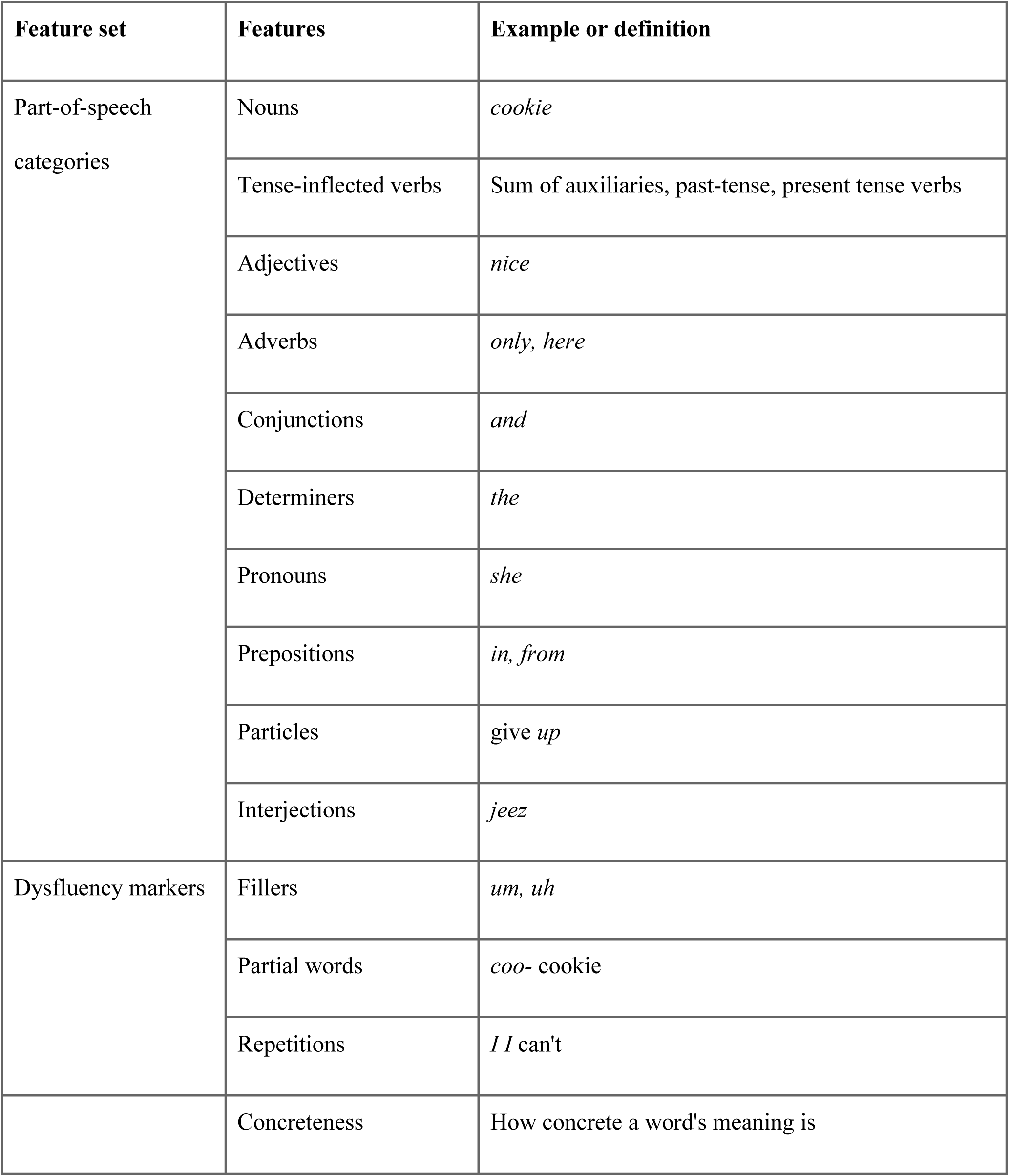
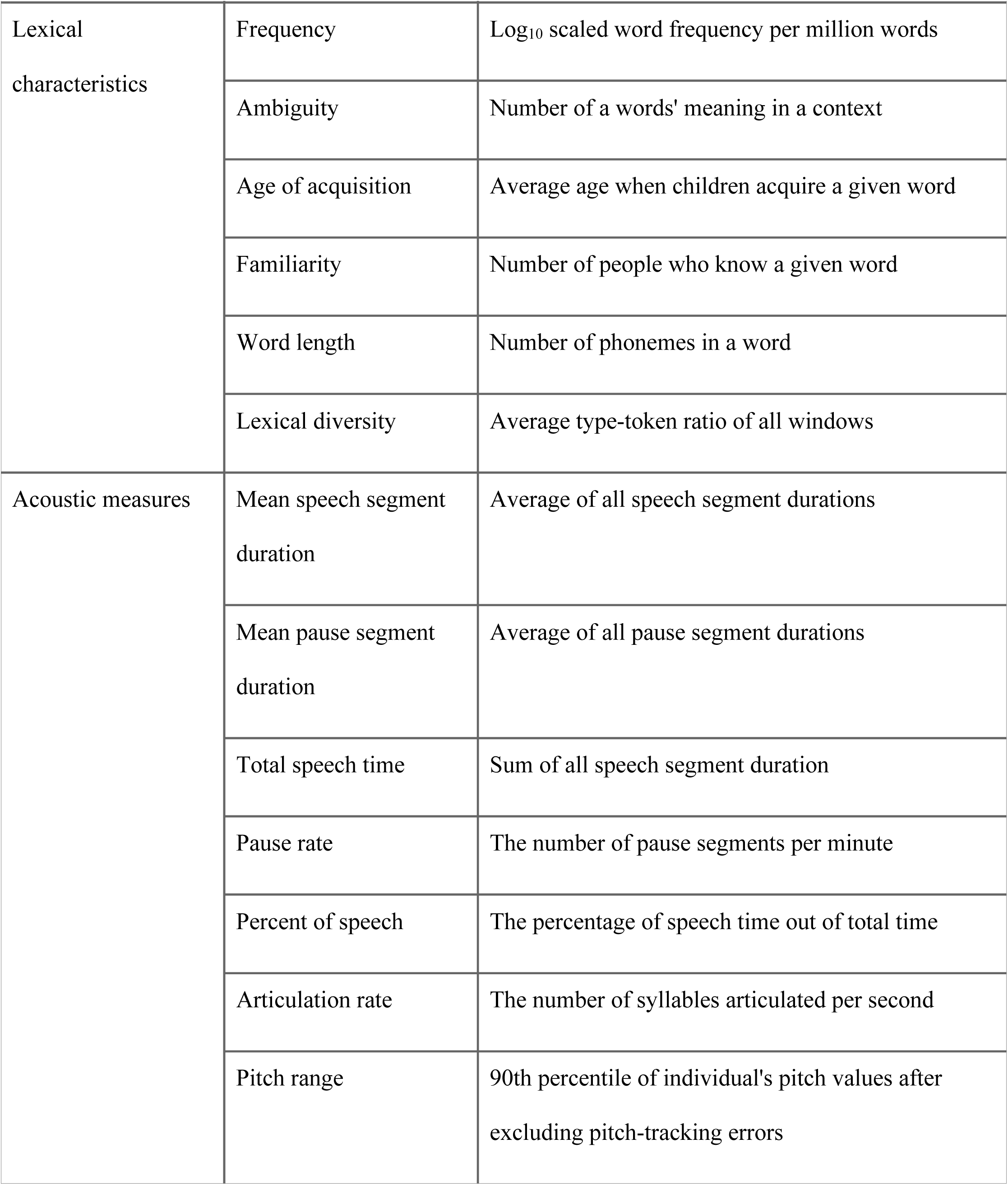
List of features used in the analysis.

